# Positive islet autoantibodies confirm classical type 1 diabetes in patients treated as type 2 diabetes who have early progression to insulin

**DOI:** 10.64898/2026.11.05.26367089

**Authors:** V.E Simpson, N Thomas, A.V Hill, J Knupp, T.J McDonald, L.A. Donelly, K.A Patel, J Murray Leech, B.M Shields, E.R Pearson, A.G Jones

## Abstract

**Objective:** Early progression to insulin following initial management as type 2 diabetes may indicate missed type 1 diabetes. Recent guidelines advise islet-autoantibody testing in all adults who progress to insulin within 3 years, though this has not been directly tested. We aimed to determine whether positive islet autoantibodies in this setting confirm the genetic characteristics and C-peptide progression of classical type 1 diabetes.

**Research Design and Methods:** We compared genetic susceptibility to type 1 and type 2 diabetes (T1DGRS and T2DGRS) and prospectively assessed C-peptide loss over 3 years after diagnosis among islet-autoantibody-positive adults initially treated as having type 2 diabetes (without insulin) who progressed to insulin within 3 years (n=86), versus participants with classical type 1 (n=499) and type 2 (n=603) diabetes. Results were replicated in cross-sectional data from the DARE/GoDARTS studies (n=9466).

**Results:** Participants who progressed to insulin within 3 years and had positive islet autoantibodies had the genetic characteristics and C-peptide loss of classical type 1 diabetes: T1DGRS −0.04 (−0.27, 0.18) vs 0.00 (−0.09, 0.09) [p=0.71], T2DGRS −0.35 (−0.59, −0.11) vs −0.35 (−0.44, −0.26) [p=0.99] and annual change in urine C-peptide creatinine ratio −47% (−55%, −40%) vs −49% (−52%, −46%) [p=0.6] respectively. In contrast, autoantibody-positive participants without early insulin (n=51) had genetic and C-peptide characteristics consistent with type 2 diabetes. Findings were similar in cross-sectional cohorts.

**Conclusion:** Islet-autoantibody-positive individuals with early insulin requirement following initial treatment as type 2 diabetes have similar genotype and progression to classical type 1 diabetes. This supports recent guidance that they should be diagnosed and treated as type 1 diabetes.

**Graphical Abstract:** 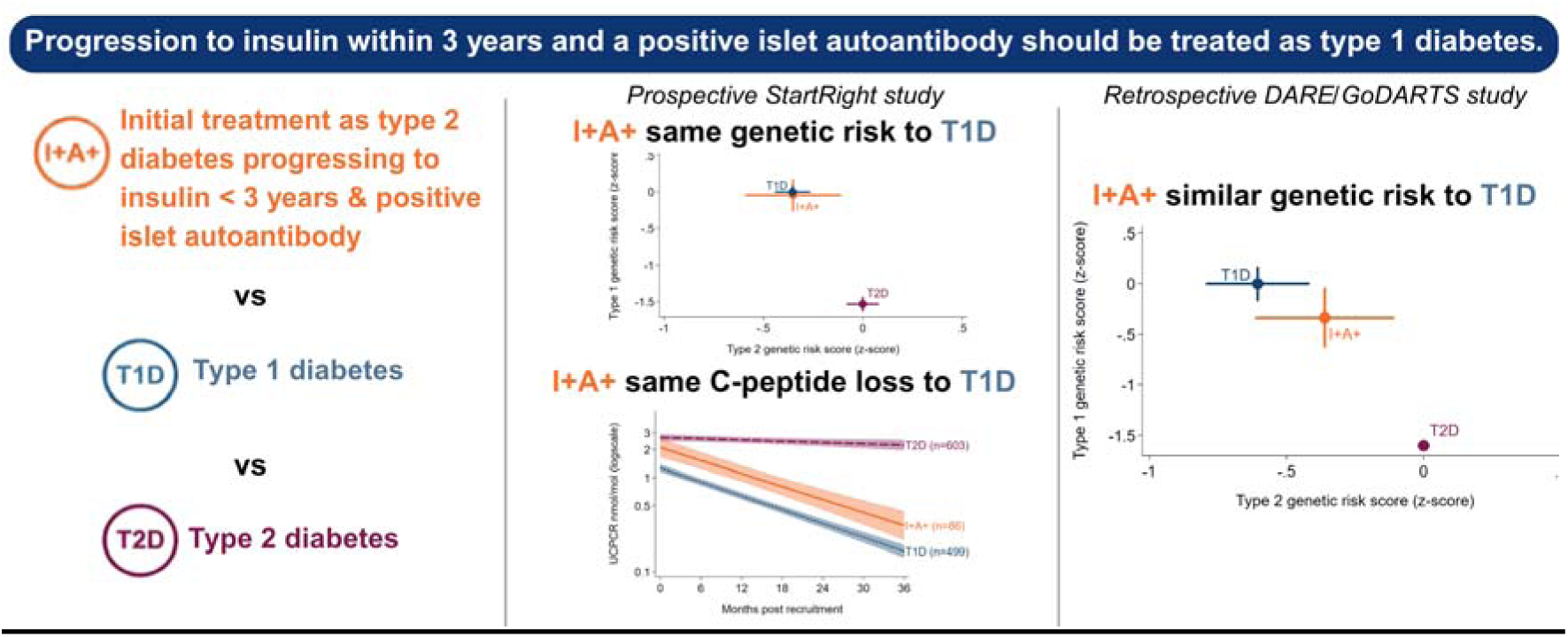

**Article highlights:** *Why did we undertake this study?:* Recent international clinical guidelines have suggested that patients initially thought to have type 2 diabetes who progress to insulin within 3 years should have islet antibody testing, and that in this setting, positive islet autoantibodies will confirm classical type 1 diabetes. The guidelines are based on the mathematical interpretation of islet-autoantibody test performance and have not been directly tested.

*What is the specific question(s) we wanted to answer?:* Does a positive islet autoantibody test in people initially treated as having type 2 diabetes (without insulin) who progress to insulin within 3 years of diagnosis identify those with the genetic characteristics and C-peptide loss seen in classical type 1 diabetes?

*What did we find?:* In prospective analysis from diagnosis, we found that individuals initially treated as type 2 diabetes who progressed to insulin within 3 years of diagnosis and had one or more positive islet autoantibodies had similar genetic susceptibility to type 1 diabetes, and rate of C-peptide loss over 3 years, as individuals with classical type 1 diabetes. In contrast, the genetic susceptibility and rate of C-peptide loss in participants autoantibody positive without insulin requirement were largely similar to antibody-negative type 2 diabetes. Findings were similar in the analysis of two cross-sectional studies.

*What are the implications of our findings?:* Our results support routine islet autoantibody testing in patients initially thought to have type 2 diabetes who require insulin within 3 years and suggest that individuals who test positive should be classified and managed as classical type 1 diabetes, in line with recent international guidelines.

## Introduction

Diagnosing type 1 diabetes in adults can be challenging as features overlap with type 2 diabetes. Even highly specific features for type 1 diabetes may have limited predictive value due to the high prevalence of type 2 diabetes (1,2). Consequently, a large proportion (∼30-40%) of those developing classical type 1 diabetes as adults are misclassified and inadvertently diagnosed and treated as type 2 diabetes (3–5).

Islet autoantibodies are the primary tests for differentiating diabetes subtypes at diagnosis (6–9). However, these tests have imperfect specificity, and when autoimmune aetiology of diabetes is unlikely (low pre-test probability, such as in those with clinical features suggesting type 2 diabetes), a single positive autoantibody may be insufficiently specific to confirm type 1 diabetes (7,10–12). In the population with positive islet autoantibodies there will therefore be those with true positive results (autoimmune aetiology diabetes) and false positive (or disease irrelevant) results (nonautoimmune (type 2) diabetes), which may explain the heterogeneous and overlapping phenotype of adults initially treated as type 2 diabetes who have positive islet autoantibodies, termed Latent Autoimmune Diabetes in Adults (LADA) (1,10,12).

Recent clinical guidelines from the American Diabetes Association (ADA) and European Association for the Study of Diabetes (EASD), as well as other organisations, have recommended that autoantibody tests be routinely performed in all patients with clinically suspected type 1 diabetes and in people initially treated as having type 2 diabetes who progress to insulin treatment within 3 years of diagnosis (8,13,14). The recommendation is based on the finding that rapid progression to insulin treatment is highly predictive of missed type 1 diabetes, and in this (high pre-test probability) setting positive islet autoantibodies are mathematically likely to be highly predictive of autoimmune aetiology diabetes (2,3,10).

We hypothesise that one or more positive islet autoantibodies, assessed using modern high-specificity assays, will confirm the presence of autoimmune diabetes in adults initially treated as type 2 diabetes who progress to insulin treatment within 3 years of diabetes diagnosis. Therefore, this patient group will have the genetic and C-peptide progression characteristics of classical type 1 diabetes. In contrast, those without early insulin (a very low pre-test probability setting) will largely consist of false positive results and have characteristics similar to type 2 diabetes. We aimed to test this hypothesis in a prospective study of newly diagnosed adult-onset diabetes, with additional replication in cross-sectional cohorts.

## Research Design and Methods

We compared C-peptide progression and genetic susceptibility to type 1 diabetes (T1DGRS) and type 2 diabetes (T2DGRS) in participants initially treated as type 2 diabetes (without insulin for over 4 weeks from diagnosis), followed prospectively for >3 years from diabetes diagnosis, and categorised according to islet autoantibody and progression to insulin status as follows:

- progressing to insulin within 3 years of diabetes diagnosis with ≥1 positive islet autoantibody (I+A+; n=86)
- progressing to insulin within 3 years of diagnosis with negative islet autoantibodies (I+A-; n=44)
- not progressing to insulin within 3 years of diagnosis with a positive islet autoantibody (I-A+; n=51).

These participants were compared to type 1 and type 2 reference groups from the same prospective study: a type 1 diabetes reference group defined by a clinical diagnosis of type 1 diabetes at study recruitment, ≥1 positive islet autoantibody and treatment with insulin within four weeks of diagnosis (n=499), and a type 2 diabetes reference group defined by islet autoantibody negativity and no progression to insulin within 3 years of diabetes diagnosis (n=603).

Findings were replicated in a combined cross-sectional cohort from two unselected population studies: DARE and GoDARTS (n=9,466).

### Prospective cohort

Adults (≥18 years) with newly diagnosed diabetes (≤12 months diabetes duration, excluding secondary or gestational diabetes) were recruited from 55 National Health Service (NHS) sites across the UK between August 2016 and February 2020 in the StartRight study (https://clinicaltrials.gov/study/NCT03737799). Participants were recruited through routine clinical care appointments and record screening in primary care and self-referral after advertisements. Individuals were followed annually for 3 years. To ensure sufficient participants with late-onset type 1 diabetes, recruitment of those aged >50 was enriched for participants receiving insulin treatment at recruitment. A study flow diagram is presented in Supplementary Fig. 2s. Participants were excluded if they received insulin within 4 weeks, but did not have a clinical diagnosis of T1D or were islet autoantibody negative. Participants who did not progress to insulin treatment within 3 years of diagnosis were excluded if they had insufficient follow up (<36 months).

At recruitment, participants self-reported diabetes type and details of initial presentation, including initial treatment and where applicable timing of insulin initiation. A research nurse confirmed the diagnosis details based on medical notes and laboratory records. A non-fasted (within 1–5 h of a meal) blood sample was taken for analysis of C-peptide, islet autoantibodies (glutamic acid decarboxylase antibody [GADA], zinc transporter 8 [ZNT8A], islet antigen 2 [IA-2A]), and DNA extracted for the generation of type 1 and type 2 diabetes genetic risk scores (T1DGRS and T2DGRS). Baseline BMI was calculated from measured weight and height.

At recruitment and at each annual visit, participants provided a urine sample in boric acid preservative for measurement of the urine C-peptide–to–creatinine ratio (UCPCR) within 2 hours of completing a home meal (15,16). After recruitment, participants were contacted annually to report their current treatment, any changes in treatment, and the date of each change. The research team obtained post-recruitment HbA1c results from healthcare records. At ≥3 years from recruitment (final visit), blood or capillary C-peptide was also assessed within 1-5 hours of a meal.

### Cross-sectional cohort

Findings were replicated in participants with a clinical diagnosis of T2D in the GoDARTS (Genetics of Diabetes Audit and Research Tayside) and DARE (Diabetes Alliance for Research in England) studies, limited to those with age of onset ≥18 years and no insulin requirement for 4 weeks. The group was compared to participants diagnosed as type 1 diabetes at age ≥18 with positive autoantibodies (GADA, IA-2A or ZNT8A) and insulin from diagnosis (DARE cohort). A study flow diagram is presented in Supplementary Fig. 7s.

GoDARTS is a cross-sectional study of individuals with a clinical diagnosis of type 2 diabetes recruited in Tayside (Scotland) between 1^st^ October 1997 and 1^st^ July 2006 (17), with post recruitment follow up from electronic healthcare records. DARE is a cross-sectional study based in the Southwest of England of an unselected population of adults with diabetes recruited between 5^th^ November 2009 and 31^st^ July 2017 (3).

In GoDARTS and DARE, clinical information was assessed by self-report and examination through an interview with a research nurse. A non-fasted blood sample was collected for C-peptide (DARE only), islet autoantibodies (DARE: GADA, IA-2A and ZNT8A; GoDARTS: GADA only), and genotyping as previously described (3,17). Progression to insulin treatment was based on self-report in the DARE cohort, and assessment of linked electronic prescription records in GoDARTS.

### Laboratory analysis

All biochemical analyses other than HbA1c values were undertaken at the Royal Devon and Exeter Hospital academic blood sciences department in Exeter, UK. HbA1c results at recruitment were analysed at local NHS laboratories. GADA, IA-2A and ZNT8A were measured using the RSR ELISA assay (RSR, Cardiff, UK) using the DYNEX DS2 automated ELISA system (DYNEX Technologies, Worthing, U.K.). Islet autoantibodies were positive if the result was higher than the 97.5^th^ centile of 1,559 control subject population without diabetes; GADA ≥11 units/mL, IA-2A ≥7.5 units/mL, and ZNT8A ≥65 units/mL in those aged <30 years and ≥10 units/mL in those aged ≥30 years (18,19). Exeter Hospital’s academic blood science department achieved 99% specificity across all three islet autoantibodies, with 74% sensitivity for GADA and ZNT8A, and 72% for IA-2A, in the 2020 international islet autoantibody standardisation program.

The Roche Diagnostics E170 immunoanalyser (Mannheim, Germany) was used to measure plasma, capillary and urine C-peptide, with a limit of detection of 3.3 pmol/L for plasma and urine, and 200 pmol/L for capillary. Inter- and intra-assay coefficients of variation were <4.5% and <3.3%, respectively. Urine creatinine was analysed with the Jaffe method on the Roche P800 modular analyser for UCPCR.

### Diabetes Genetic Risk Scores

We generated the T1DGRS from 67 single-nucleotide polymorphisms associated with type 1 diabetes, accounting for interactions among 18 HLA-DR-DQ haplotype combinations as previously reported (20). LGC Genomics genotyped single-nucleotide polymorphisms (21). A T2DGRS was calculated using a previously validated risk score obtained from the PGS Catalog (PGS002720) (22). Standardised GRS are presented (T1DGRS standardised to type 1 diabetes controls and T2DGRS standardised to type 2 diabetes controls).

### Reporting of Results to Participants and Clinicians

In the StartRight study, islet autoantibody results and brief advice (see Supplementary Figure 1s) were provided to participants and their treating clinicians after the baseline visit. Participants and treating clinicians were unaware of the baseline or follow-up C-peptide, or genetic results. The individual’s treatment was left to the responsible clinician. In GoDARTS and DARE, autoantibody, C-peptide and genetic results were not fed back to participants.

### Statistical Analysis

Mean type 1 and type 2 genetic risk scores and 95% CIs were presented for each insulin/antibody subgroup and compared to the reference type 1 and type 2 diabetes groups using t-tests.

Annual rate of loss of C-peptide (StartRight only), measured by UCPCR (assessed annually from recruitment to 3 years) was compared between subgroups (I+A+, I+A-, I-A+) and the two reference groups (type 1 diabetes and type 2 diabetes) using a mixed-effects model of UCPCR (23), with time since recruitment (in months, grouped into 6-month intervals and restricted to the first 36 months), subgroup and a subgroup*time interaction as fixed effects, and a random intercept and random slope for time for each individual. UCPCR data were natural log transformed (ln) for analysis due to skewness, allowing a linear fit, in line with previous studies (23–26). Differences in slopes between the key subgroups (I+A+, I+A-, I-A+) and the two reference groups (type 1 diabetes and type 2 diabetes) were assessed by using the subgroup*time interaction terms in the models. Coefficients are presented as annual percentage change per year by back-transforming the fixed-effect coefficient to the original scale and multiplying by 100 [(exponential(12 x β-coefficient) – 1)*100], where β is the monthly log scale slope. 95% confidence intervals were derived using the same equation on the log-scale confidence intervals. The half-life of UCPCR was determined from ln(0.5)/β, representing the time in months for UCPCR to decline by 50%.

Blood (plasma/capillary) C-peptide at recruitment and at the ≥3 year follow-up visit (measured on two occasions only) in StartRight is reported as median (IQR), and as the percentage below 200 pmol/L (severe insulin deficiency). The proportion of severe insulin deficiency was assessed using chi-squared. Slope for blood C-peptide (measured at baseline and final visit) was not assessed because the lower limit of the C-peptide assay when assessed by capillary sample is 200 pmol/L (76% [969/1,283] of those with follow up samples), making this less sensitive for determining slopes compared to UCPCR and biasing comparisons. C-peptide analysis in DARE reported median (IQR) C-peptide levels at recruitment, limited to those with diabetes duration >3 years at C-peptide assessment, and compared proportions <200 pmol/L (DARE only) using chi-squared.

Secondary analysis included limiting analysis to those progressing to insulin after six months of diabetes duration (in line with the widely used definition of LADA) (27,28) and exploration of whether GAD titre and the number of islet autoantibodies (1 vs >1 positive autoantibody) were associated with genetic risk and C-peptide progression in antibody-positive participants with and without early progression to insulin using the same approaches described above. Consistent with a previous study, GAD titres were log-transformed and classified using a two-component Gaussian finite mixture model (11). GAD titres were classified as high or low based on the nadir of this model in StartRight (602 units/mL in StartRight), with the same GAD titre cut-off used in the DARE/GoDARTS analysis.

Data are reported as mean (SD) unless otherwise specified. All analysis was conducted in StataSE 18.0 (StataCorp, College Station, TX).

## Results

Overall participant characteristics (prospective StartRight study) are presented in Table 1, and the flowchart for participant selection is shown in Supplementary Figure 2s (StartRight). At recruitment, when islet autoantibodies were measured, the median duration of diabetes was 4.9 months (IQR 2-9). The median diabetes duration at the latest follow-up (with blood C-peptide measurement) was 52 months (IQR 45-60). Of the individuals initially treated as having type 2 diabetes (without insulin), 130 progressed to insulin within 3 years from diagnosis (median (IQR) time to insulin initiation 6 months (2–15)), of which 66% (86/130) were islet autoantibody-positive (59% of these (51/86) were multiple antibody positive). 654 individuals did not progress to insulin within 3 years of diagnosis of whom 8% (51/654) were islet autoantibody-positive; 10% of these (5/51) were multiple antibody-positive.

**Table 1:** baseline characteristics (StartRight) of subgroups, defined by islet autoantibody status and progression to insulin. Mean with standard deviation are given for continuous variables unless otherwise stated. I +/−indicates progression to insulin between 4 weeks and 3 years and islet antibody status (A +/−). T1D – reference type 1 diabetes cohort (clinical diagnosis T1D at recruitment, autoantibody positive, insulin within 4 weeks of diagnosis). T2D – reference T2D cohort (clinical diagnosis T2D, autoantibody negative, no insulin treatment for >3 years post diagnosis), * at diabetes presentation/diagnosis **Median 4.9 months (IQR 2 − 9) diabetes duration, ***median 52 (IQR 47, 56) months diabetes duration, note due to missing data for follow up C-peptide denominator shown for % <200 pmol/L.

| StartRight | I+A+ (n =86) | I-A+ (n =51) | I+A- (n =44) | T1D (n =499) | T2D (n =603) |
|---|---|---|---|---|---|
| **Patient factors** |  |  |  |  |  |
| Age at Diagnosis years (SD) | 45.6 (13.9) | 50.1 (10.3) | 46.8 (14.3) | 37.3 (14.0) | 51.0 (12.7) |
| Female %(n) | 36%(31/86) | 41%(21/51) | 23%(10/44) | 40%(200/499) | 41%(249/603) |
| BMI kg/m2 (SD) | 25.2 (4.3) | 33.3 (6.9) | 29.0 (7.7) | 25.0 (4.4) | 32.9 (6.8) |
| White ethnicity %(n) | 93%(80/86) | 94%(48/51) | 84%(37/44) | 96%(479/499) | 91%(549/603) |
| Co-existent autoimmune disease %(n) | 23%(20/86) | 20%(10/51) | 5%(2/44) | 15%(76/499) | 7%(44/603) |
| Osmotic symptoms* %(n) | 78%(67/86) | 73%(37/51) | 80%(35/44) | 96%(479/499) | 59%(358/603) |
| Unintentional weight loss* %(n) | 67%(58/86) | 14%(7/51) | 43%(19/44) | 86%(430/499) | 16%(94/603) |
| Months to start insulin Median (IQR) | 6.0 (2.3, 11.6) |  | 8.3 (3.2, 22.1) | 0.0 (0.0, 0.0) |  |
| **Biochemistry** |  |  |  |  |  |
| Random glucose* mmol/L (SD) | 17.1 (6.7) | 11.6 (6.0) | 15.8 (8.1) | 21.4 (9.4) | 11.8 (7.1) |
| HbA1c* mmol/mol (SD) | 88 (25) | 71 (24) | 95 (31) | 106 (27) | 68 (24) |
| HbA1c* % (SD) | 10.2 (2.3) | 8.6 (2.2) | 10.8 (2.8) | 11.9 (2.5) | 8.4 (2.2) |
| HDL** mmol/L (SD) | 1.30 (0.51) | 1.30 (1.13) | 1.10 (0.32) | 1.42 (0.56) | 1.19 (0.56) |
| LDL** mmol/L (SD) | 2.87 (0.99) | 3.21 (1.32) | 3.45 (1.19) | 2.91 (1.04) | 2.99 (1.07) |
| Triglycerides** mmol/L Median (IQR) | 1.70 (1.10, 2.30) | 2.00 (1.50, 2.80) | 2.40 (1.60, 5.20) | 1.40 (0.90, 2.40) | 2.00 (1.40, 3.00) |
| **Post-meal blood C-peptide** |  |  |  |  |  |
| Recruitment** pmol/L Median (IQR) | 656 (376, 1080) | 1930 (1410, 2980) | 1425 (803, 2085) | 474 (281, 694) | 1860 (1380, 2610) |
| >3 years post diagnosis*** pmol/L Median (IQR) | 199 (143, 506) | 2033 (1166, 2670) | 1280 (605, 1639) | 199 (73, 298) | 1683 (1243, 2370) |
| **C-peptide categories (>3 years)** |  |  |  |  |  |
| <200 pmol/L %(n) | 55%(34/62) | 0%(0/46) | 10%(2/21) | 62%(212/341) | <1%(2/499) |

### Individuals with early progression to insulin and a positive islet autoantibody have the genetic characteristics of classical type 1 diabetes

Figure 1 shows that individuals who progressed to insulin and had a positive islet autoantibody (I+A+) had similar type 1 diabetes genetic susceptibility (assessed by T1DGRS) to those with antibody-positive classical type 1 diabetes: mean (95%CI) T1DGRS −0.04 (−0.27,0.18) vs 0.00 (−0.09,0.09) respectively [p=0.71]. Type 2 diabetes genetic susceptibility was also similar in these two groups: T2DGRS −0.35 (−0.59, −0.11) vs −0.35 (−0.44, −0.26) [p=0.99]. In comparison to confirmed type 2 diabetes, I+A+ individuals had higher T1DGRS −0.04 (−0.27,0.18) vs −1.53 (−1.63,-1.43) [p<0.001] and lower T2DGRS −0.35 (−0.59,-0.11) vs 0.00 (−0.08,0.08) [p=0.01].

**Figure 1:**
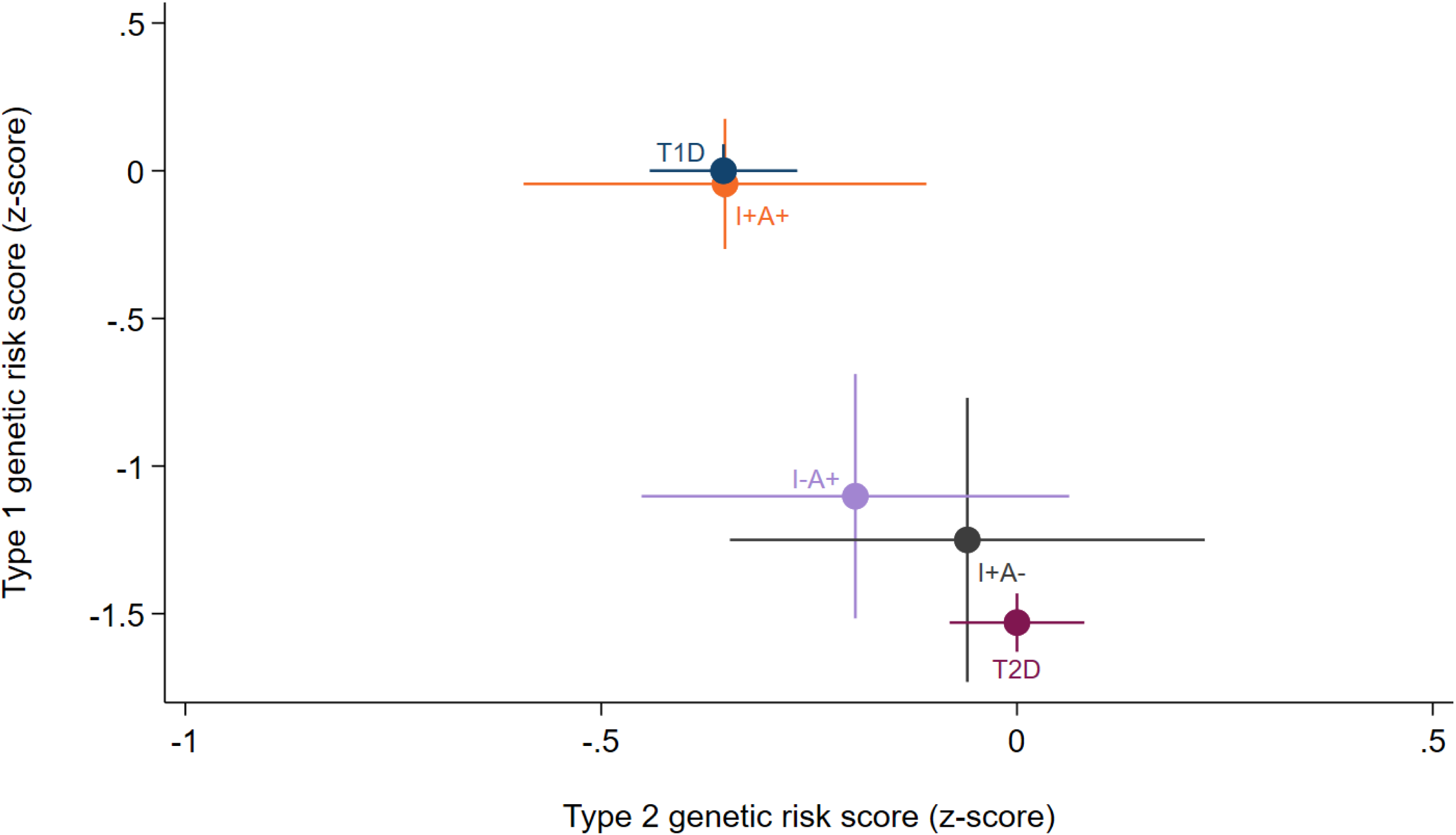
Type 1 and type 2 genetic risk scores (StartRight) for prospectively studied individuals who were islet autoantibody positive or negative and either did or did not progress to insulin compared to a reference group of individuals with type 1 diabetes and type 2 diabetes. Progression to insulin (I+) between 4 weeks and 3 years or no progression (I-), islet autoantibody status (A+/−). T1D – reference type 1 diabetes cohort (clinical diagnosis T1D at recruitment, autoantibody positive, insulin within 4 weeks of diagnosis). T2D – reference T2D cohort (clinical diagnosis T2D, autoantibody negative, no insulin treatment for >3 years post diagnosis) Error bars represent 95% confidence interval.

In contrast, participants who did not progress to insulin but were islet autoantibody-positive (I-A+) had similar type 2 diabetes genetic susceptibility to classical type 2 diabetes, and only minor enrichment for type 1 diabetes genetic risk: T1DGRS (I-A+ vs type 2 diabetes) −1.10 (−1.52,-0.69) vs −1.53 (−1.63,-1.43) [p=0.05] and T2DGRS − 0.19 (−0.45,0.06) vs 0.00 (−0.08,0.08) [p=0.15], see Figure 1.

### Participants who progressed to insulin and had positive islet autoantibodies have a comparable rate of C-peptide decline to classical type 1 diabetes

Annualised rate of change in UCPCR, and baseline and follow-up blood C-peptide levels, are shown in Figure 2 and Table 1 respectively. C-peptide loss, assessed by UCPCR, was similar in I+A+ compared to type 1 diabetes; mean (95%CI) annual UCPCR change −47% (−55%,-40%) with a half-life of 13 months (11,17) vs −49% (−52%,-46%) and a half-life of 12 months (11,13) [p=0.6] respectively. T2D had slower C-peptide progression (−6% (−11%, −1%)), half-life 133 months (73,622) [p (vs I+A+) <0.001]. The proportion of individuals who developed blood C-peptide <200 pmol/L at latest follow-up (median 4.1 years post diagnosis) was similar between I+A+ and T1D: 55% (34/62) vs 62% (212/341), respectively [p=0.37], and substantially higher than type 2 diabetes [<1% (2/499), p<0.001].

**Figure 2:**
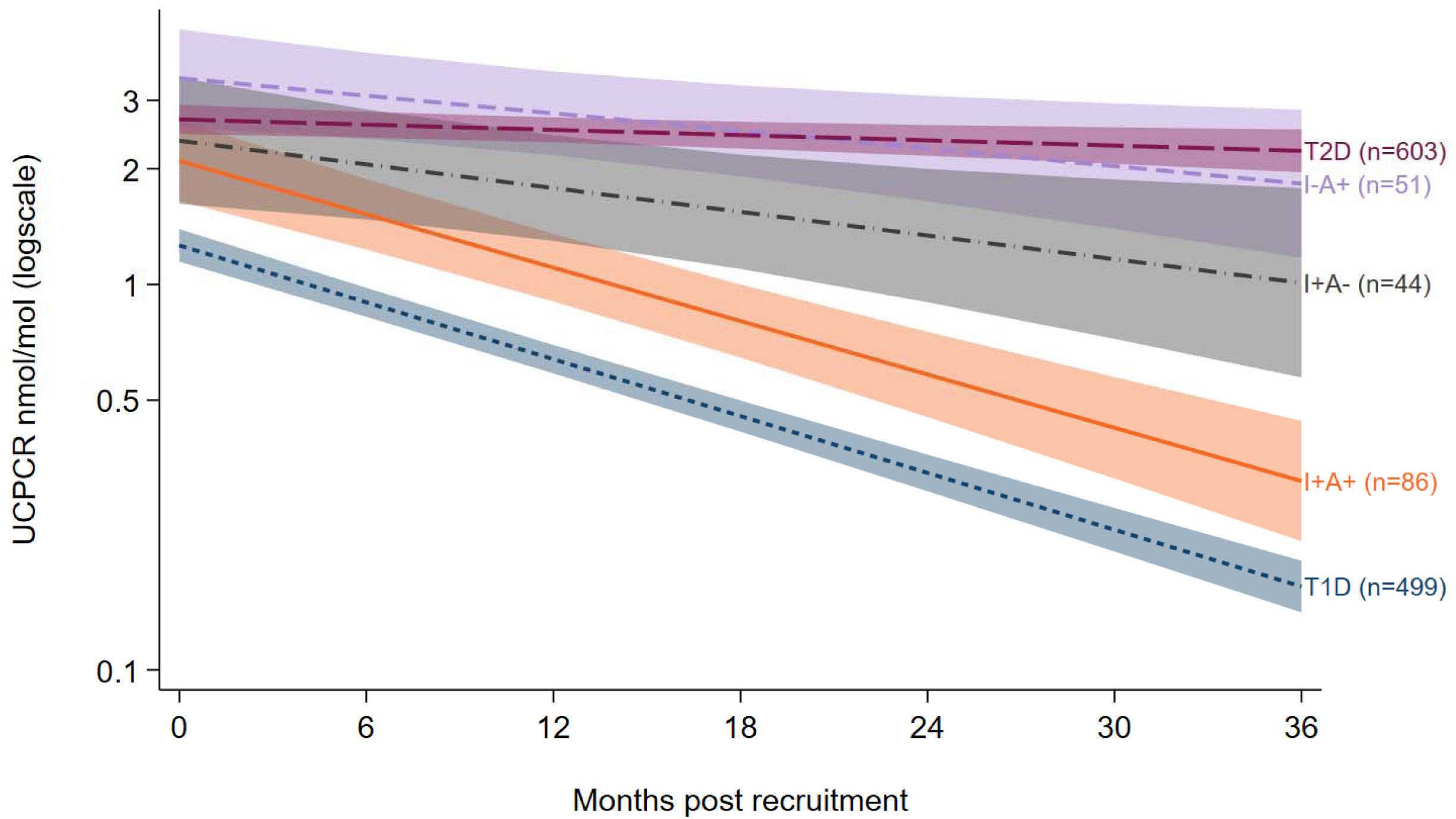
Urine C-peptide creatinine ratio [UCPCR] (StartRight) change from recruitment for groups defined by islet autoantibody status and progression to insulin. Lines show predicted UCPCR from the mixed-effects model (see Statistical Analysis), plotted on a log scale with axis values back-transformed to nmol/mol. Shaded areas represent 95% confidence intervals. Progression to insulin (I+) between 4 weeks and 3 years or no progression (I-), islet autoantibody status (A+/−). T1D – reference type 1 diabetes cohort (clinical diagnosis T1D at recruitment, autoantibody positive, insulin within 4 weeks of diagnosis). T2D – reference T2D cohort (clinical diagnosis T2D, autoantibody negative, no insulin treatment for >3 years post diagnosis)

Although rates of C-peptide loss did not differ between I+A+ and type 1 diabetes, median blood C-peptide and UCPCR at recruitment were higher in I+A+ compared to type 1 diabetes (median serum C-peptide 656 (376,1080) pmol/L vs 474 (281,694) pmol/L [p<0.001] and UCPCR 2.0 (0.9, 2.8) vs 1.1 (0.6, 1.8) nmol/mmol [p<0.001]) but lower than type 2 diabetes (1860 (1380,2610) pmol/L [p<0.001] and 3.5 (3.3,3.8) nmol/mmol [p<0.001]) (Table 1).

In autoantibody positive individuals not progressing to insulin (I-A+) C-peptide loss was substantially lower than type 1 diabetes and did not significantly differ from type 2 (mean annual change UCPCR −19% (−33%,-5%) and half-life of 40 months (21,162) [p vs T1D <0.001, vs T2D p=0.63]. None of the I-A+ participants developed C-peptide <200 pmol/L.

### Restricting analysis to those progressing to insulin after 6 months did not meaningfully change findings

Sensitivity analysis restricted to those progressing to insulin after 6 months of diagnosis (I+A+ n=43) did not alter findings (Supplementary Figure 3s), with similar genetic risk and UCPCR loss in the I+A+ subgroup, again comparable with T1D [p >0.3 for all]. C-peptide <200 pmol/L at follow-up remained similar to individuals with classical type 1 diabetes [48% vs 62%, p=0.29] (Supplementary Table 1s).

### Antibody number and titre were not associated with type 1 and 2 diabetes genetic susceptibility in autoantibody-positive patients with early progression to insulin

In antibody-positive participants with early progression to insulin (I+A+), 41% (35/86) were single autoantibody-positive, of whom the majority (83% (29/35)) were GADA-positive. In the I+A+ group, antibody number was not associated with an altered type 1 or 2 diabetes genetic susceptibility (Single vs multiple antibody positive T1DGRS − 0.07 (−0.47,0.33) vs −0.03 (−0.29,0.23) [p=0.86], T2DGRS −0.51 (−0.87,-0.14) vs −0.25 (−0.58,0.08) [p=0.29]), see Supplementary Figure 4s. Among I+A+ participants who were GADA-positive, GADA titre showed no relationship with genetic susceptibility or UCPCR progression (Supplementary Figures 5s and 6s).

In contrast, while there was no relationship between GADA titre and C-peptide progression, I+A+ participants with single positive autoantibodies had lower loss of C-peptide than those with multiple positive autoantibodies: UCPCR annual change (annual change: −29% (−44%,-14%) [n=35] vs −58% (−66%,-50%) [n=51], p<0.001;

Supplementary Figure 4s). Severe insulin deficiency at >3 years was also less frequent in single-antibody-positive individuals (32%; 8/25) than in multi-antibody-positive individuals (70%; 26/37) [p=0.03] (Supplementary Table 2s).

### Replication of findings in an independent cohort of individuals with longstanding diabetes

Results were replicated in participants from the independent DARE/GoDARTS cross-sectional cohorts (Supplementary Figure 7s). In those diagnosed as type 2 diabetes and treated without insulin for at least 4 weeks, GADA autoantibodies and C-peptide were measured at median 7.6 (IQR 4.4-12.2) years after diabetes diagnosis. 391 participants progressed to insulin between 4 weeks and 3 years, of which 20% (77/391) were autoantibody positive (median time to insulin 18 months; IQR 10-26).

The relationship between subgroup and both type 1 and type 2 genetic risk in DARE/GoDARTS is shown in Figure 3. Consistent with the prospective analysis, T1DGRS was markedly raised and T2DGRS low in those autoantibody positive with early insulin treatment (I+A+). However, in contrast to the prospective study, T1DGRS was modestly reduced in comparison to participants with classical type 1 diabetes and insulin from diagnosis (n = 147): T1DGRS −0.33 (−0.63,-0.04) vs 0.00 (−0.17,0.17) [p=0.04]. This difference appeared to be driven by lower T1DGRS in those proceeding to insulin between 2 and 3 years after diagnosis, compared with those with more rapid progression to insulin, where T1DGRS was consistent with type 1 diabetes (Supplementary Figure 8s).

**Figure 3:**
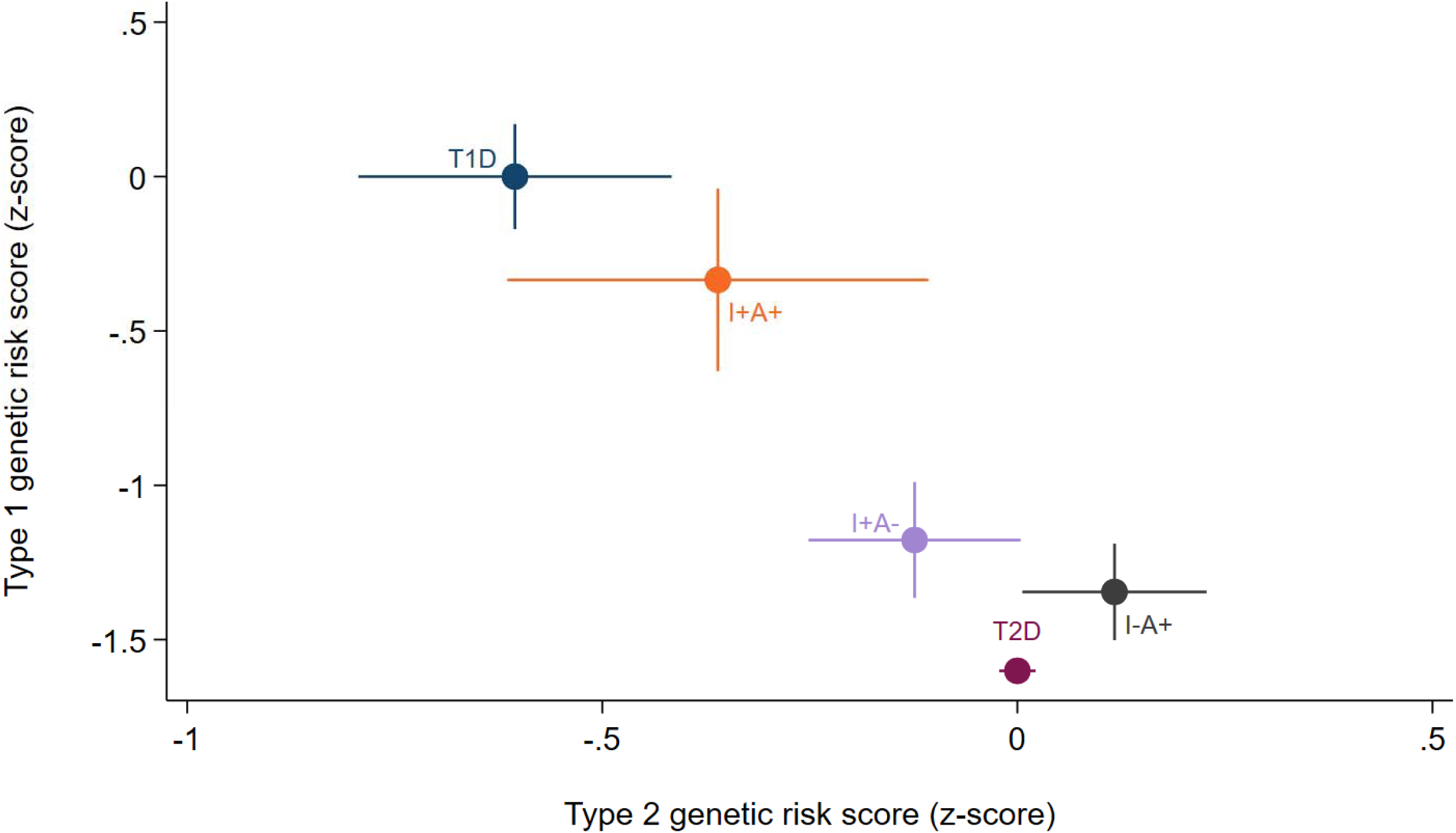
Type 1 and 2 genetic risk scores for individuals in the DARE/GoDARTS cohorts, split by islet autoantibody status and progression to insulin, compared with a reference group of individuals with type 1 diabetes and type 2 diabetes. Progression to insulin (I+) between 4 weeks and 3 years or no progression (I-), islet autoantibody status (A+/−). T1D – reference type 1 diabetes cohort (DARE participants with a clinical diagnosis of T1D at age ≥18 years, autoantibody positive (GADA, IA-2A or ZNT8A), insulin from diagnosis). T2D – reference T2D cohort (clinical diagnosis T2D at age ≥18 years, autoantibody negative, no insulin treatment for >3 years post diagnosis). Error bars represent 95% confidence intervals

DARE/GoDARTS participants with positive autoantibodies without early progression to insulin (I-A+) had the type 1 and 2 diabetes genetic characteristics consistent with classical type 2 diabetes, without evidence of enrichment for the characteristics of T1D (Figure 3 and Supplementary Table 3s).

When measured at a median of 14.2 (IQR 7.3-21.6) years of diabetes duration, random serum C-peptide levels (DARE only) were low in the vast majority of I+A+ cases, with a median C-peptide of 23 (3-200)pmol/L (n=39) and 74% having C-peptide <200pmol/L.

In GADA-positive individuals with rapid progression to insulin, GADA titre was not associated with genetic susceptibility in DARE/GoDARTS (Supplementary Figure 9s).

## Discussion

We have shown that positive islet autoantibodies are associated with the genetic and C-peptide progression characteristics of classical type 1 diabetes in individuals who progress to insulin rapidly (within 3 years of diabetes diagnosis), suggesting a diagnosis of type 1 diabetes is usually appropriate, in line with recent international guidelines. In contrast, most individuals who were islet autoantibody-positive but did not progress to insulin had genetic characteristics and C-peptide loss similar to classical autoantibody-negative type 2 diabetes. Findings were largely consistent in cross sectional data from those with longstanding clinically diagnosed type 2 diabetes.

A key strength of the StartRight study is the routine testing of islet autoantibodies at diagnosis and C-peptide in newly diagnosed adults with diabetes, with follow up for more than 3 years after diagnosis. The systematic collection of islet autoantibody status allows us to assess the utility of this test without biasing results by who was tested, and prospective study allows robust assessment of decline in C-peptide and progression to insulin. Another key strength is the inclusion of GRS, as clinicians are blinded to the result. Therefore, GRS can be used as an independent marker of diabetes type within a cohort, whereas clinical features will reflect the diagnosis to which a patient is assigned (29). Replication in a completely independent cross-sectional cohort, assessed using the same islet autoantibody and C-peptide assays in the same laboratory, and the same assessment of genetic susceptibility, provides reassurance that the findings are robust.

A limitation of our analysis in the StartRight study is that autoantibody status was fed back to the treating clinician team, which may have influenced the decision to start insulin. However, this applies to all autoantibody positive participants, and if clinicians were starting insulin unnecessarily based on islet autoantibody results, we would expect this to bias results of the I+A+ groups towards a type 2 diabetes phenotype, resulting in (for example) slower loss of endogenous insulin secretion than classical type 1 diabetes in this group. In contrast, we found that this group did not differ from classical type 1 diabetes, suggesting this is unlikely to be the case. Importantly, many autoantibody-positive individuals never progressed to insulin, suggesting that clinicians predominantly relied on disease progression or other clinical features rather than autoantibody status alone to initiate insulin. Similar findings were observed in DARE/GoDARTS, which did not feedback autoantibody results. Further limitations include that the moderate numbers of islet autoantibody-positive individuals found among those initially treated as type 2 diabetes in prospective and cross-sectional cohorts mean confidence intervals are wide in subgroup analyses, and that autoantibodies in DARE/GoDARTS were limited to GADA and measured many years after diabetes diagnosis.

Our findings are consistent with previous research that has shown an association between insulin requirement and the phenotypic and/or genetic characteristics of type 1 diabetes in those diagnosed with type 2 diabetes who are autoantibody positive (30–32). For example, Radtke et al., using data from the HUNT study, showed that patients meeting LADA criteria, who did not require insulin at a median of 8.1 ± 0.9 years of follow-up, had similar C-peptide levels and other clinical characteristics to autoantibody-negative type 2 diabetes (32), which is consistent with our findings for autoantibody positive individuals without insulin requirement at 3 years. Our findings of little or no association between autoantibody titre and both C-peptide progression, and type 1 diabetes genetic susceptibility are largely consistent with previous studies in clinically diagnosed type 1 diabetes (11,33). However, they contrast with the majority of studies in the overall population of individuals with apparent type 2 diabetes, in which higher titres and numbers have been associated with a more type 1-like phenotype and genotype (27,34). This difference, as we have previously reported, may be explained by Bayes’ theorem (1,10). In high-prior-probability settings (clinically diagnosed type 1 diabetes, or rapid progression to insulin) even a single positive autoantibody is likely to confirm autoimmune diabetes. Here, factors that further increase test specificity (higher titre, more antibodies) will not meaningfully further increase test positive predictive value. In low-prior-probability settings (clinically suspected type 2 diabetes) a single positive autoantibody will be insufficiently specific to confirm autoimmune aetiology diabetes: and a mix of true and false positives is expected. Factors that increase test specificity (such as higher antibody titre or number) result in a test positive population with a higher proportion of true autoimmune diabetes (higher positive predictive value), and therefore a more type 1 diabetes like phenotype.

### Clinical implications

Our results support recent ADA/EASD and UK type 1 diabetes immunotherapy consortium guidelines, suggesting autoantibody testing for all who progress to insulin within 3 years, even when they are thought to have type 2 diabetes, with a positive test (1 or more positive autoantibodies) usually confirming type 1 diabetes (8,14). Our results suggest that it is usually appropriate to treat this group of patients as classical type 1 diabetes, even if only one autoantibody is positive. These individuals will usually have a similar aetiology, loss of insulin secretion (and therefore treatment requirements) to autoantibody-positive adults who are recognised as having type 1 diabetes at diagnosis. In contrast, our results suggest that where patients initially thought to have type 2 diabetes are positive for a single autoantibody, the majority of those who do not require insulin within 3 years are likely to have nonautoimmune aetiology type 2 diabetes. However, most current guidelines consider all patients with a positive autoantibody to have a form of autoimmune type 1 diabetes, despite the imperfect specificity of these assays (35,36). A caution in interpreting our findings, and a key area for further research, is that we do not have sufficient sample size to determine whether a 3-year cut-off, recommended in EASD/ADA guidelines and supported by previous research, is optimal (3,8). Although in small numbers, our preliminary analysis suggests that the predictive value of positive autoantibodies will be high but (in the absence of other supporting evidence of type 1 diabetes) imperfect in those progressing to insulin within 2 and 3 years from diabetes diagnosis.

In conclusion, islet-autoantibody-positive individuals with early insulin requirement following initial treatment for type 2 diabetes have similar genotype and progression to classical type 1 diabetes. This supports recent international guidance that these patients should be diagnosed and treated as type 1 diabetes.

## Authors and contributions

V.E.S., N.J.T., and A.G.J. designed the study. A.V.H., B.K., J.M.L J.K., T.J.M., ERP and A.G.J. researched the data. V.E.S., N.J.T., B.M.S., and A.G.J. analysed data. V.E.S., and A.G.J. wrote the first draft of the manuscript. All authors reviewed the draft and contributed to the revision of the manuscript.

## Data access and responsibility statement

A.G.J. is the guarantor of this work and, as such, had full access to all data in the study and takes responsibility for the integrity and accuracy of the data analysis.

## Ethics statement

The StartRight study was approved by the South West – Cornwall & Plymouth NHS Research Ethics Committee on 06/06/2016, ref: 16/SW/0130.

## Funding

The StartRight study was funded by Diabetes UK (17/0005624) and the UK National Institute of Health and Social Care Research (CS-2015-15-018). Genotyping for generation of the type 1 genetic risk score was supported by the European Foundation for the Study of Diabetes (2016 Rising Star Fellowship).

## Data Availability

Data are available upon reasonable request to the authors.

## Acknowledgement

This study was supported by the National Institute for Health and Care Research Exeter Biomedical Research Centre and National Institute for Health and Care Research Exeter Clinical Research Facility. The views expressed are those of the authors and not necessarily those of the NIHR or the Department of Health and Social Care.

## Conflict of interest statement

NT has received travel support, speaker fees and reimbursement for advisory board membership from Sanofi. All other authors declare that there are no relationships or activities that might bias, or be perceived to bias, their work.

## Prior Presentation

58th EASD Annual Meeting of the European Association for the Study of Diabetes 2022 The North European Young Diabetologists (NEYD) Annual Meeting 2023

## Notes

### Author Declarations

The StartRight study was approved by the South West (Cornwall & Plymouth) NHS Research Ethics Committee on 06/06/2016, ref: 16/SW/0130.

