## Supplementary for "Positive islet autoantibodies confirm classical type 1 diabetes in patients treated as type 2 diabetes who have early progression to insulin"

*Figure 1s: Template of information fed back to participants with positive islet autoantibody results.*

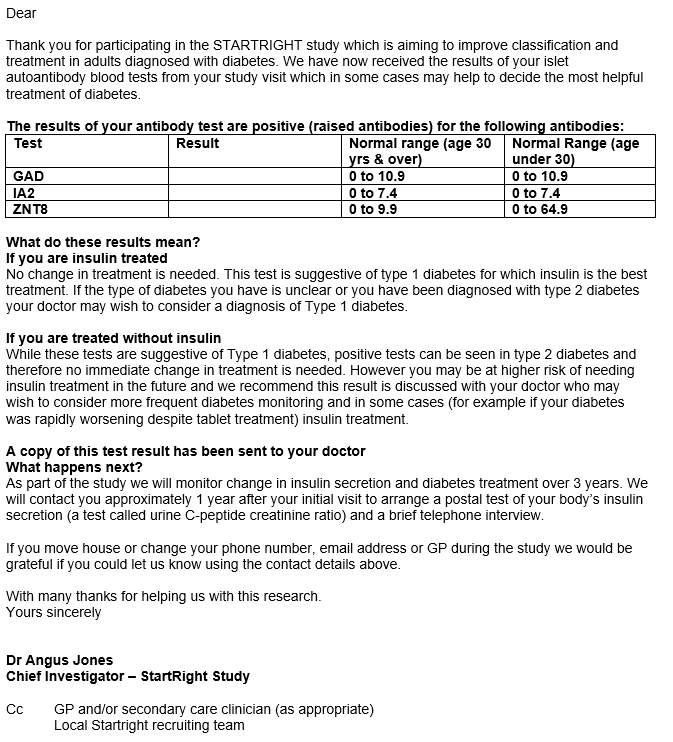

Figure 2s: Study flowchart for the StartRight study

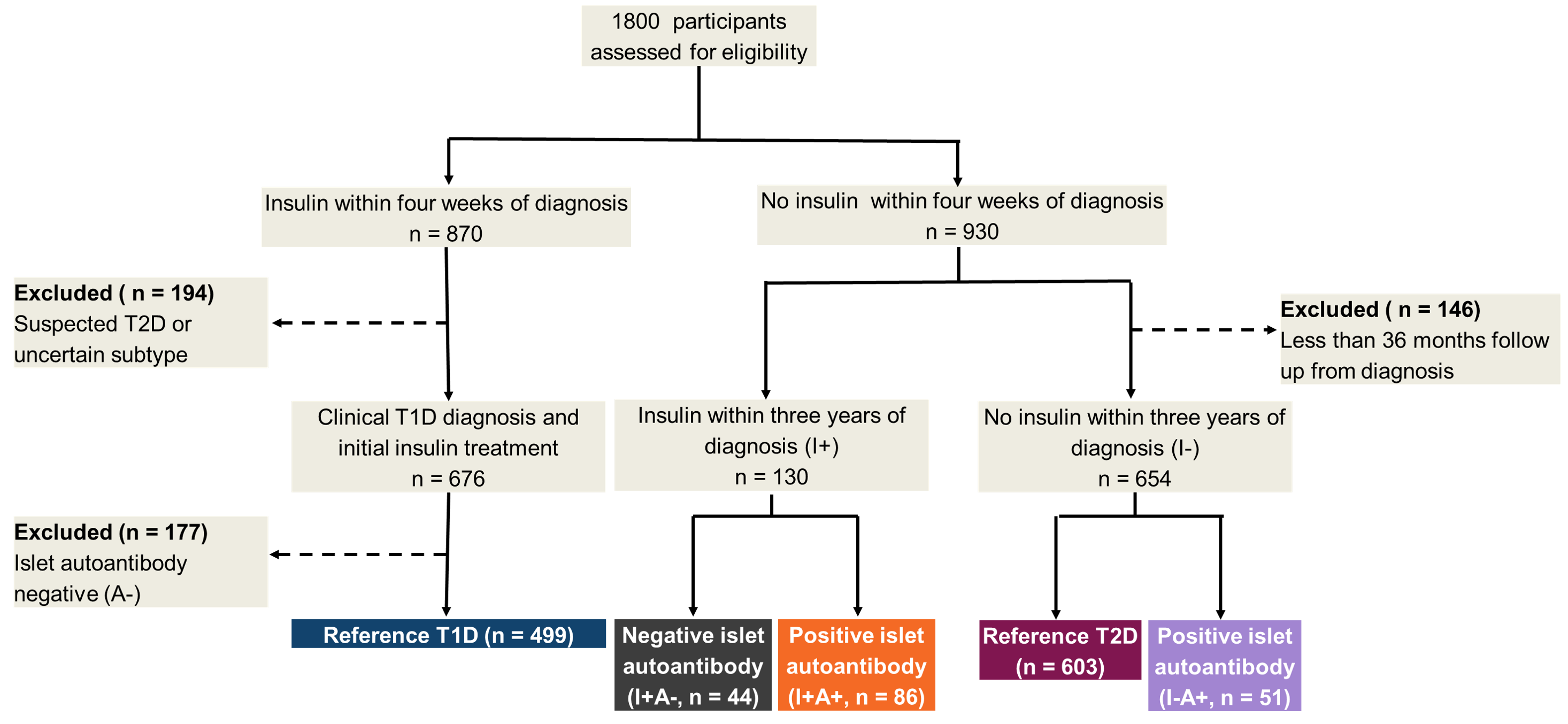

| StartRight | I+A+ (n =43) | I-A+ (n =51) | I+A- (n =25) | T1D (n =499) | T2D (n =603) |
| --- | --- | --- | --- | --- | --- |
| **Patient factors** |  |  |  |  |  |
| Age at Diagnosis years (SD) | 43.2 (13.1) | 50.1 (10.3) | 43.8 (14.6) | 37.3 (14.0) | 51.0 (12.7) |
| Female %(n) | 33%(14/43) | 41%(21/51) | 24%(6/25) | 40%(200/499) | 41%(249/603) |
| BMI kg/m2 (SD) | 24.0 (4.2) | 33.3 (6.9) | 30.8 (9.3) | 25.0 (4.4) | 32.9 (6.8) |
| White ethnicity %(n) | 88%(38/43) | 94%(48/51) | 84%(21/25) | 96%(479/499) | 91%(549/603) |
| Co-existent autoimmune disease %(n) | 21%(9/43) | 20%(10/51) | 8%(2/25) | 15%(76/499) | 7%(44/603) |
| Osmotic symptoms* %(n) | 70%(30/43) | 73%(37/51) | 76%(19/25) | 96%(479/499) | 59%(358/603) |
| Unintentional weight loss* %(n) | 65%(28/43) | 14%(7/51) | 40%(10/25) | 86%(430/499) | 16%(94/603) |
| Months to start insulin Median (IQR) | 11.6 (7.5, 18.7) |  | 19.1 (11.6, 32.7) | 0.0 (0.0, 0.0) |  |
| **Biochemistry** |  |  |  |  |  |
| Random glucose* mmol/L (SD) | 14.6 (6.7) | 11.6 (6.0) | 14.2 (7.6) | 21.4 (9.4) | 11.8 (7.1) |
| HbA1c* mmol/mol (SD) | 84 (27) | 71 (24) | 83 (28) | 106 (27) | 68 (24) |
| HbA1c* % (SD) | 9.8 (2.5) | 8.6 (2.2) | 9.7 (2.6) | 11.9 (2.5) | 8.4 (2.2) |
| HDL** mmol/L (SD) | 1.38 (0.59) | 1.30 (1.13) | 1.05 (0.35) | 1.42 (0.56) | 1.19 (0.56) |
| LDL** mmol/L (SD) | 2.69 (0.80) | 3.21 (1.32) | 2.96 (0.98) | 2.91 (1.04) | 2.99 (1.07) |
| Triglycerides** mmol/L Median (IQR) | 1.40 (0.95, 2.30) | 2.00 (1.50, 2.80) | 2.10 (1.60, 5.45) | 1.40 (0.90, 2.40) | 2.00 (1.40, 3.00) |
| **Post-meal blood C-peptide** |  |  |  |  |  |
| Recruitment** pmol/L Median (IQR) | 873 (445, 1100) | 1930 (1410, 2980) | 1490 (800, 2150) | 474 (281, 694) | 1860 (1380, 2610) |
| >3 years post diagnosis*** pmol/L Median (IQR) | 228 (191, 566) | 2033 (1166, 2670) | 1370 (212, 1639) | 199 (73, 298) | 1683 (1243, 2370) |
| **C-peptide categories (>3 years)** |  |  |  |  |  |
| <200 pmol/L %(n) | 48%(15/31) | 0%(0/46) | 18%(2/11) | 62%(212/341) | 0%(2/499) |

Table 1s: baseline characteristics (StartRight) of the different groups, defined by islet autoantibody status and progression to insulin for sensitivity analysis restricting early insulin progression to between 6 months and 3 years. Mean with standard deviation are given for continuous variables. I +/- indicates progression to insulin between 6 months and 3 years and A +/- islet antibody status. T1D – reference type 1 diabetes cohort (clinical diagnosis T1D at recruitment, autoantibody positive, insulin within 4 weeks of diagnosis). T2D – reference T2D cohort (clinical diagnosis T2D, autoantibody negative, no insulin treatment for >3 years post diagnosis). * at diabetes presentation/diagnosis **Median 4.9 months (IQR 2 - 9) diabetes duration, ***median 52 (IQR 47, 56) months diabetes duration, note due to missing data for follow up C-peptide denominator shown for % <200 pmol/L.

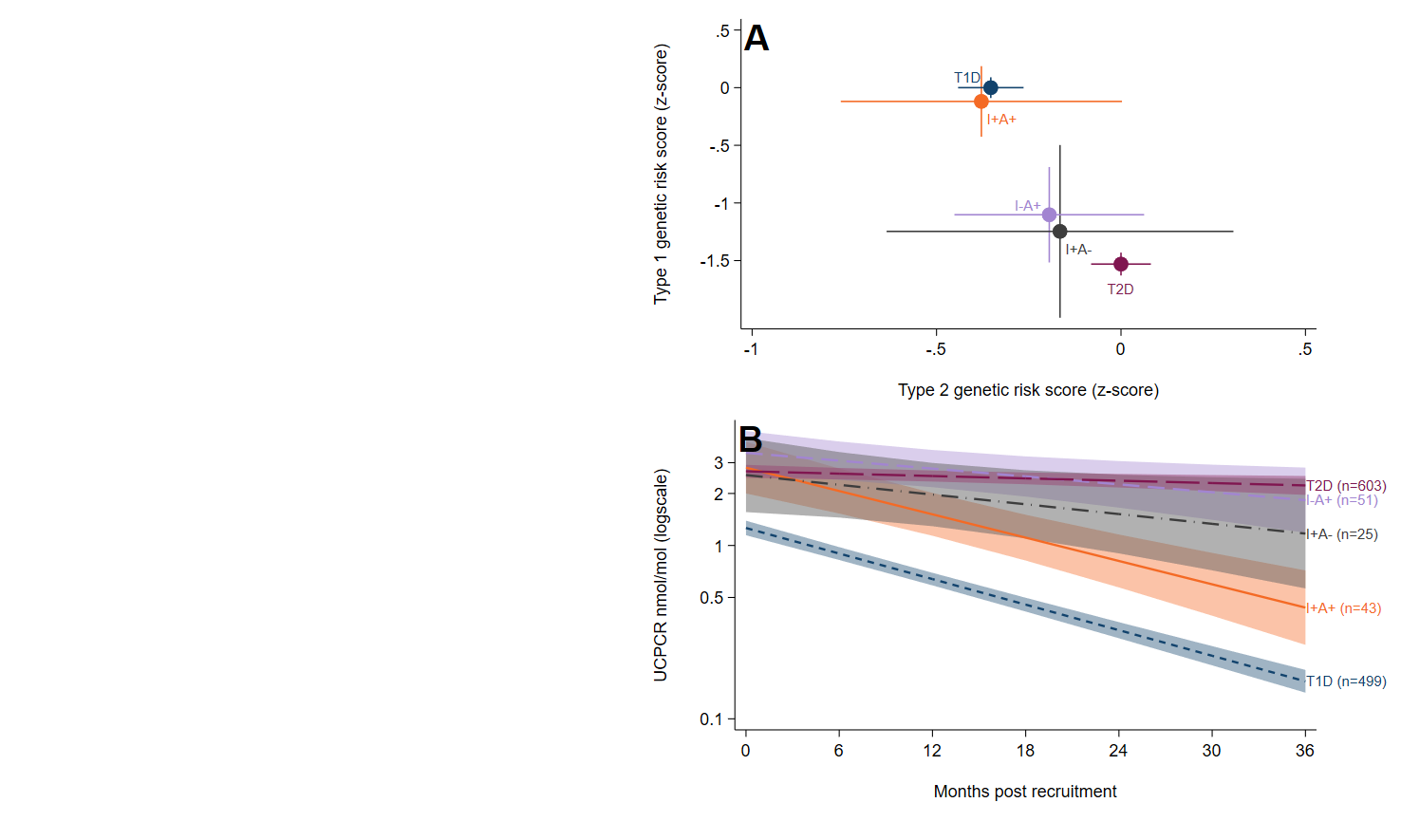

Figure 3s: Sensitivity Analysis (StartRight Study) for participants not treated with insulin within 6 months of diagnosis: genetic risk score (A) and C-peptide (UCPCR) progression (B) restricting analysis (non-type 1 diabetes subgroups only). I +/- indicates progression to insulin between 6 months and 3 years and A +/- islet antibody status. Error bars (A) and shaded areas (B) represent 95% confidence intervals.

| StartRight | Single antibody progressing to insulin  (n =35) | Multi-antibody progressing to insulin  (n =51) | T1D  (n =499) | T2D  (n =603) |
| --- | --- | --- | --- | --- |
| **Patient factors** |  |  |  |  |
| Age at Diagnosis years (SD) | 43.5 (12.3) | 47.1 (14.8) | 37.3 (14.0) | 51.0 (12.7) |
| Female %(n) | 31%(11/35) | 39%(20/51) | 40%(200/499) | 41%(249/603) |
| BMI kg/m2 (SD) | 25.6 (3.9) | 25.0 (4.5) | 25.0 (4.4) | 32.9 (6.8) |
| White ethnicity %(n) | 94%(33/35) | 92%(47/51) | 96%(479/499) | 91%(549/603) |
| Co-existent autoimmune disease %(n) | 17%(6/35) | 27%(14/51) | 15%(76/499) | 7%(44/603) |
| Osmotic symptoms* %(n) | 77%(27/35) | 78%(40/51) | 96%(479/499) | 59%(358/603) |
| Unintentional weight loss* %(n) | 71%(25/35) | 65%(33/51) | 86%(430/499) | 16%(94/603) |
| Months to start insulin Median (IQR) | 7.5 (3.6, 18.7) | 4.6 (2.1, 8.8) | 0.0 (0.0, 0.0) |  |
| **Biochemistry** |  |  |  |  |
| Random glucose* mmol/L (SD) | 17.3 (7.0) | 17.0 (6.5) | 21.4 (9.4) | 11.8 (7.1) |
| HbA1c* mmol/mol (SD) | 88 (25) | 88 (25) | 106 (27) | 68 (24) |
| HbA1c* % (SD) | 10.2 (2.3) | 10.2 (2.3) | 11.9 (2.5) | 8.4 (2.2) |
| HDL** mmol/L (SD) | 1.24 (0.38) | 1.34 (0.58) | 1.42 (0.56) | 1.19 (0.56) |
| LDL** mmol/L (SD) | 3.28 (0.98) | 2.60 (0.92) | 2.91 (1.04) | 2.99 (1.07) |
| Triglycerides** mmol/L Median (IQR) | 1.75 (1.35, 2.90) | 1.45 (0.95, 2.00) | 1.40 (0.90, 2.40) | 2.00 (1.40, 3.00) |
| **Post-meal blood C-peptide** |  |  |  |  |
| Recruitment** pmol/L Median (IQR) | 896 (435, 1100) | 584 (328, 1030) | 474 (281, 694) | 1860 (1380, 2610) |
| >3 years post diagnosis*** pmol/L Median (IQR) | 495 (199, 751) | 199 (104, 228) | 199 (73, 298) | 1683 (1243, 2370) |
| **C-peptide categories (>3 years)** |  |  |  |  |
| <200 pmol/L %(n) | 32%(8/25) | 70%(26/37) | 62%(212/341) | 0%(2/499) |

Table 2s: baseline characteristics (StartRight) of islet autoantibody-positive participants who progressed to insulin (I+A+), split by number of positive islet autoantibodies (single vs multiple), compared with reference type 1 and type 2 diabetes cohorts. Means with standard deviations are given for continuous variables. I +/- indicates progression to insulin between 4 weeks and 3 years and A +/- islet autoantibody status. T1D – reference type 1 diabetes cohort (clinical diagnosis T1D at recruitment, autoantibody positive, insulin within 4 weeks of diagnosis). T2D – reference T2D cohort (clinical diagnosis T2D, autoantibody negative, no insulin treatment for >3 years post diagnosis). * at diabetes presentation/diagnosis**Median 4.9 months (IQR 2 - 9) diabetes duration, ***median 52 (IQR 47, 56) months diabetes duration, note due to missing data for follow up C-peptide denominator shown for % <200 pmol/L.

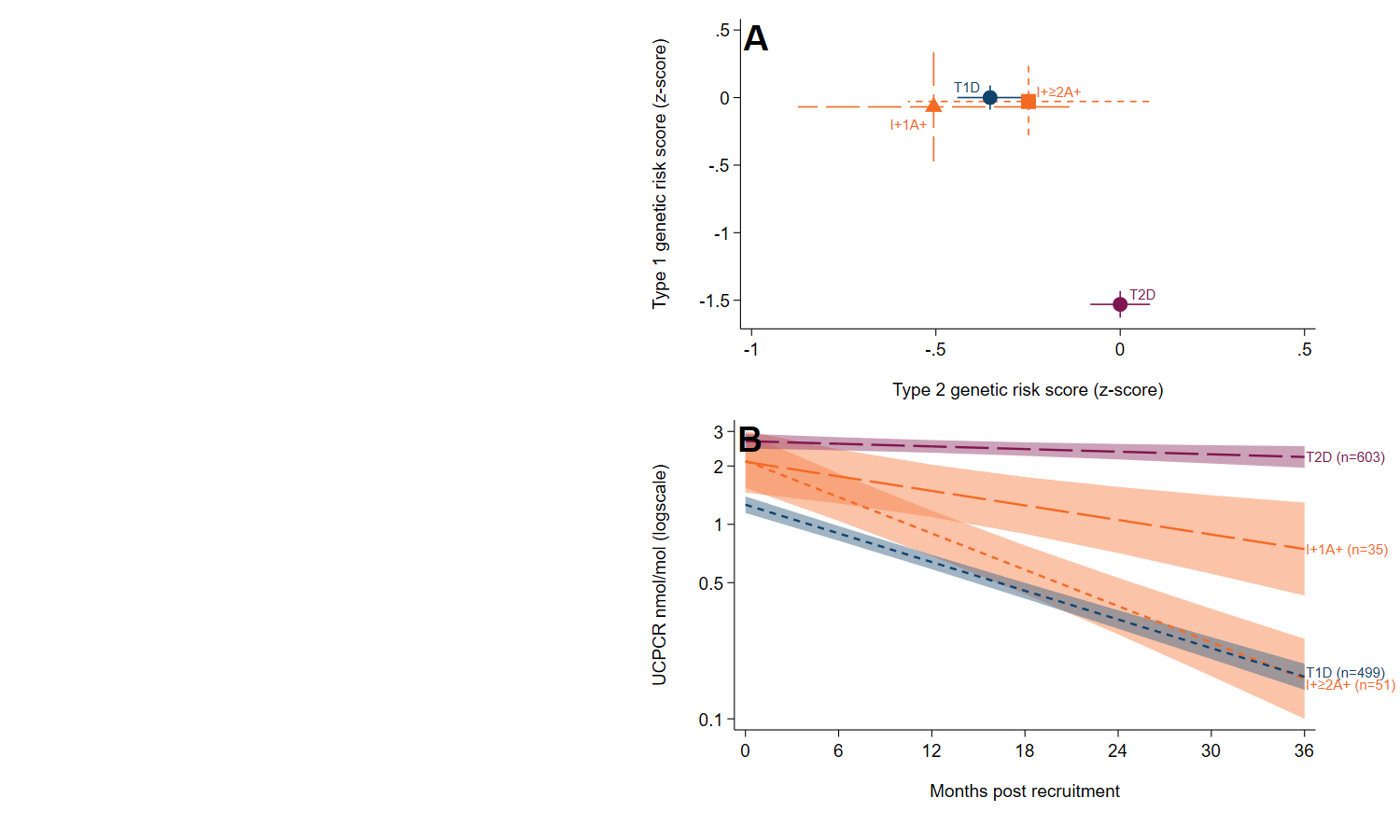

Figure 4s: Sensitivity analysis of genetic risk score (A) and C-peptide (UCPCR) progression (B) assessing the impact of autoantibody number. I +/- indicates progression to insulin between 4 weeks and 3 years, 1A+ single autoantibody positive (orange triangle), ≥2A+ two or three positive islet autoantibodies (orange square). Shaded areas represent 95% confidence intervals.

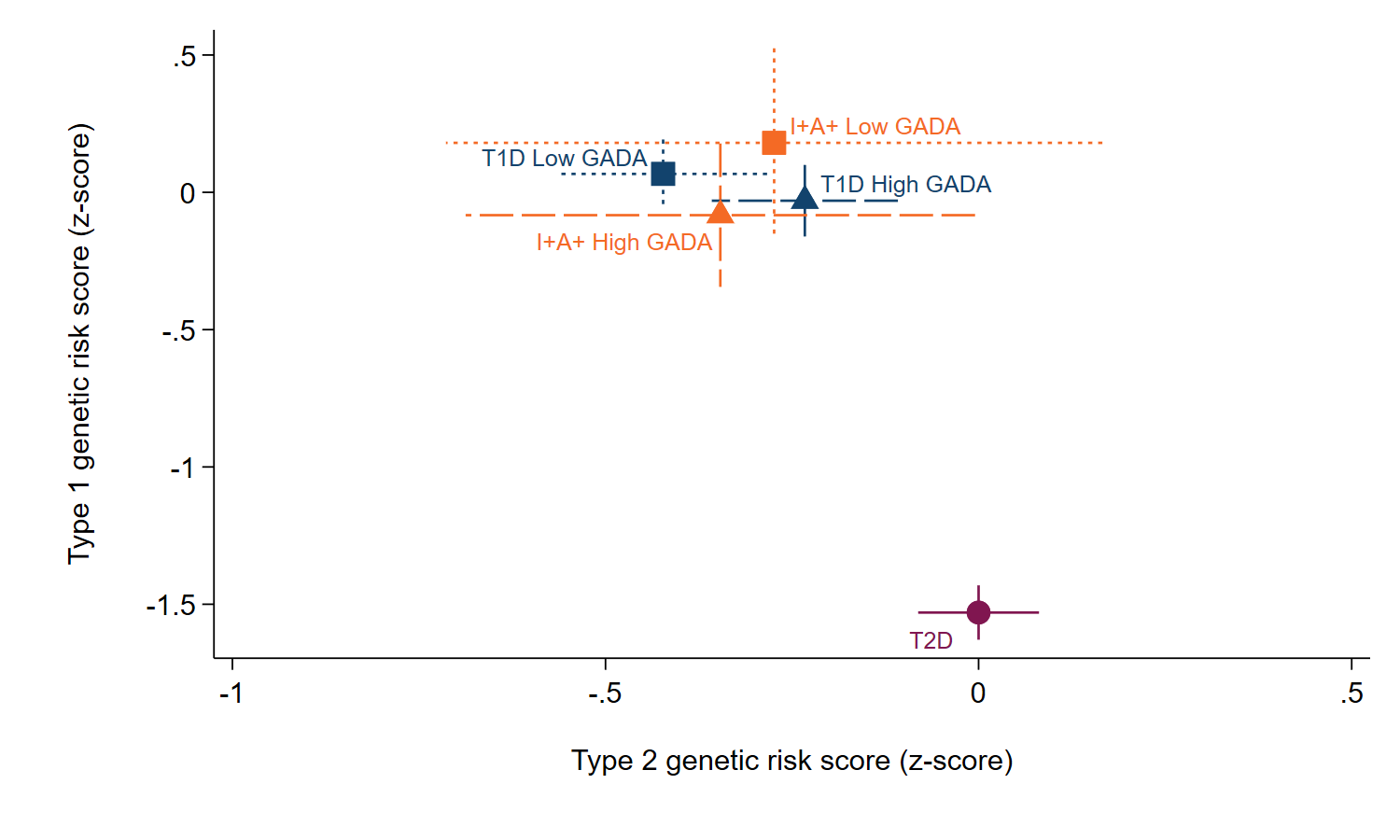

Figure 5s: Genetic risk score for individuals in StartRight who progressed to insulin (I+, 4 weeks to 3 years from diagnosis) with a positive GADA antibody (A+, orange), split by low/high GADA titre (high GADA triangle, low GADA square) and progression to insulin, compared with a reference group of individuals with type 1 diabetes (dark blue) and type 2 diabetes (purple). Error bars represent 95% confidence interval.

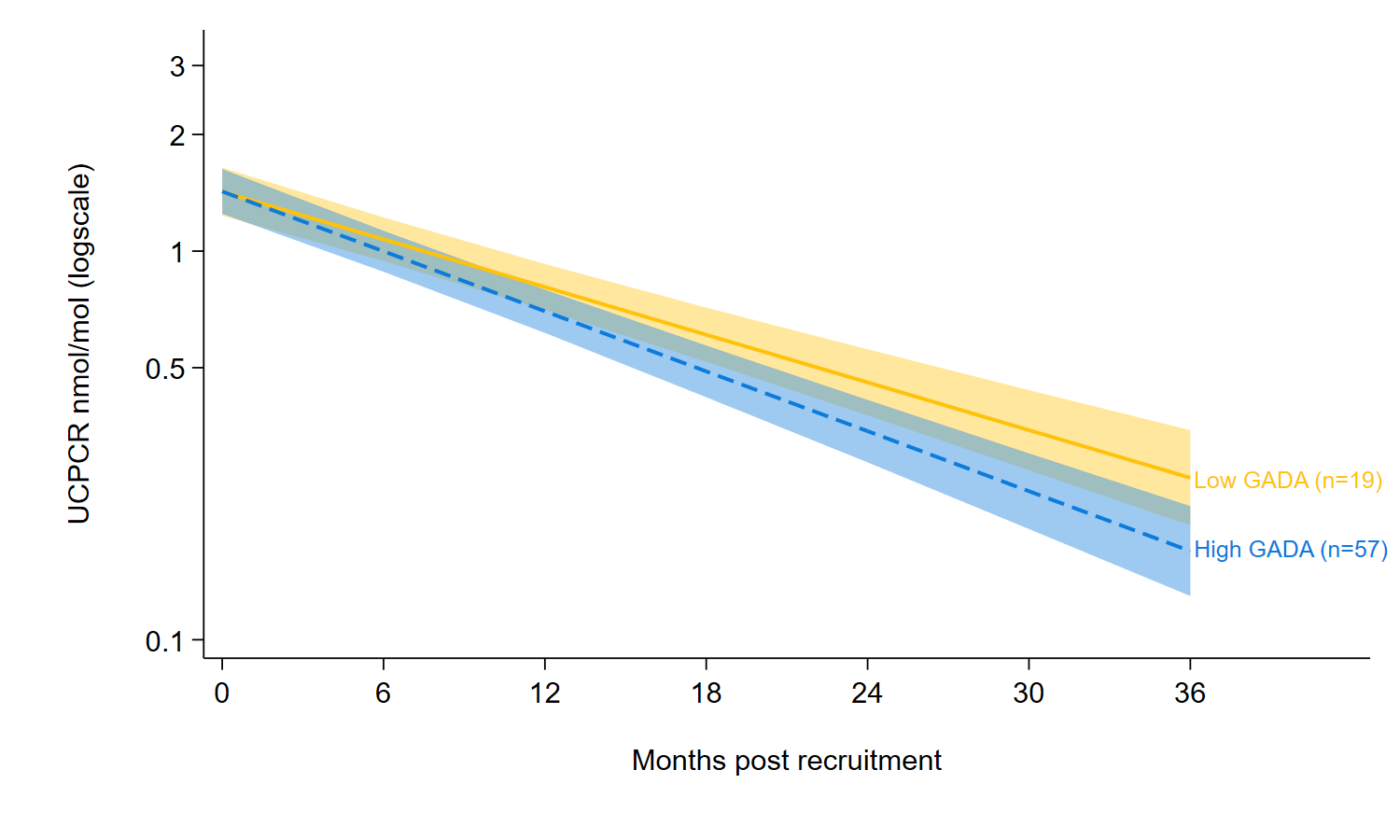

Figure 6s: Urine C-peptide creatinine ratio (UCPCR, StartRight) change from diagnosis in GADA positive individuals (88% of I+A+ group, 76/86) who progressed rapidly to insulin (4 weeks-3 years) split by low (yellow solid line) and high GADA (blue dash line) titre. Shaded areas represent 95% Confidence interval.

*Figure 7s: Study flowchart for DARE/GoDARTS study*
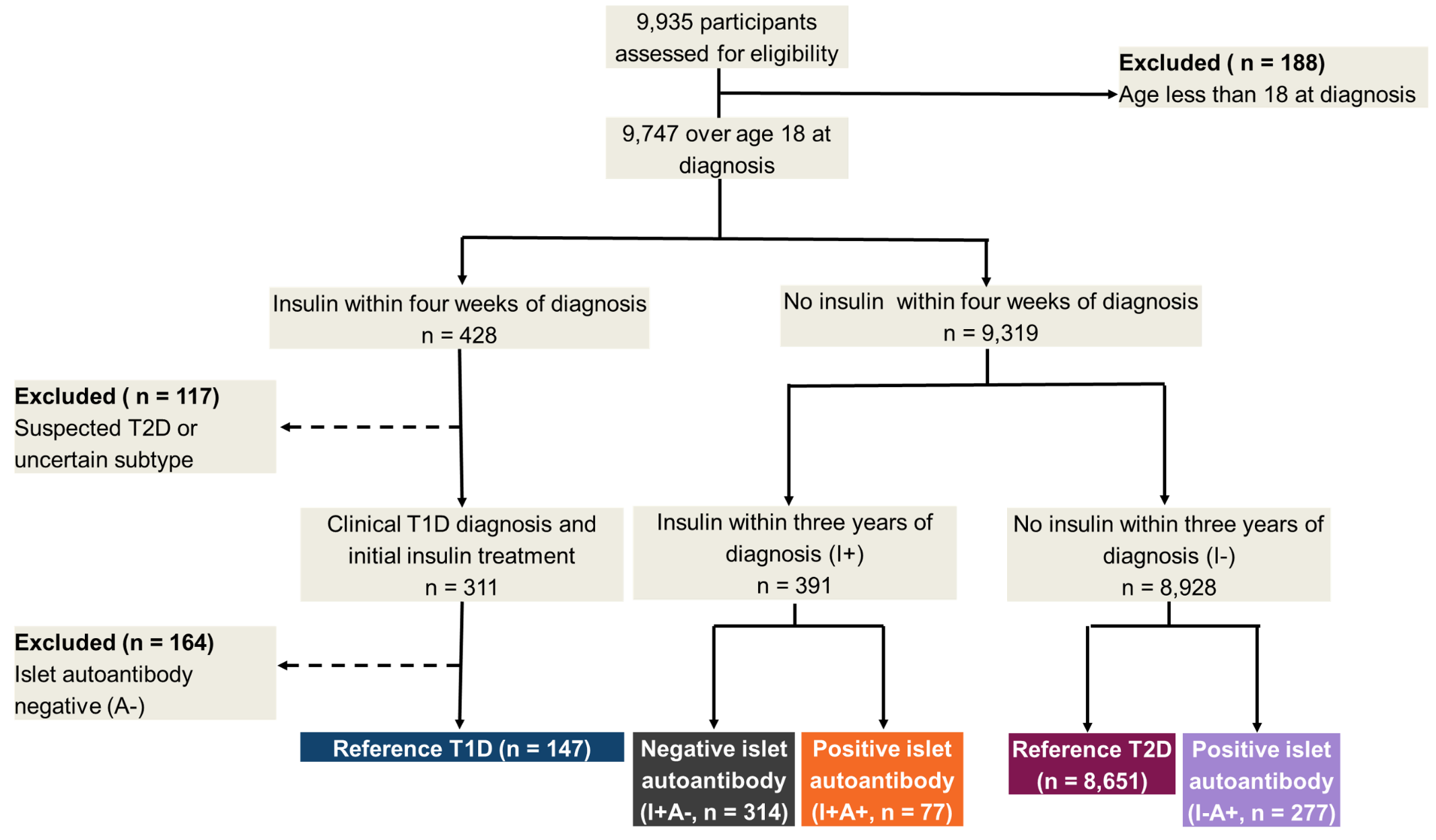

| DARE/GoDARTS | I+A+(n =77) | I-A+ (n =277) | I+A- (n =314) | T1D (n =147) | T2D (n =8651) |
| --- | --- | --- | --- | --- | --- |
| **Patient factors** |  |  |  |  |  |
| Age at Diagnosis years (SD) | 50.8 (13.8) | 59.0 (13.2) | 53.5 (12.1) | 32.9 (13.6) | 61.4 (11.3) |
| Female %(n) | 49%(38/77) | 51%(140/277) | 47%(148/314) | 49%(72/147) | 42%(3673/8651) |
| BMI kg/m^2^ (SD) | 28.5 (5.2) | 29.3 (6.2) | 31.6 (6.8) | 25.6 (4.5) | 30.5 (6.2) |
| White ethnicity %(n) | 100%(77/77) | 99%(275/277) | 100%(313/314) | 97%(143/147) | 99%(8580/8651) |
| Months to start insulin Median (IQR) | 18.0 (10.0, 26.0) |  | 24.0 (12.0, 29.0) | 0.0 (0.0, 0.0) |  |
| **Biochemistry at diagnosis** |  |  |  |  |  |
| HbA1c mmol/mol (SD) | 82 (23) | 65 (25) | 81 (27) | 77 (30) | 61 (21) |
| HbA1c % (SD) | 9.7 (2.1) | 8.1 (2.3) | 9.5 (2.4) | 9.2 (2.7) | 7.8 (1.9) |
| **C-peptide at recruitment** |  |  |  |  |  |
| Duration at C-peptide measurement months Median (IQR) | 170.2 (87.7, 259.8) | 148.5 (83.4, 275.4) | 195.0 (125.5, 271.8) | 112.2 (47.8, 314.5) | 108.8 (56.7, 188.9) |
| C-peptide pmol/L Median (IQR) | 23 (3, 200) | 630 (105, 1665) | 742 (436, 1350) | 9 (3, 67) | 1240 (766, 2050) |
| **C-peptide categories (at recruitment)** |  |  |  |  |  |
| <200 pmol/L %(n) | 74%(29/39) | 26%(11/43) | 19%(19/100) | 92%(95/103) | 2%(15/718) |

Table 3s: Characteristics of the DARE/GoDARTS cohorts, split by progression to insulin between 4 weeks and 3 years and antibody status. Progression to insulin (I+) between 4 weeks and 3 years or no progression (I-), islet autoantibody status (A+/-). T1D – reference type 1 diabetes cohort (DARE participants with a clinical diagnosis of T1D at age ≥18 years, autoantibody positive (GADA, IA-2A or ZNT8A), insulin from diagnosis). T2D – reference T2D cohort (clinical diagnosis T2D at age ≥18 years, autoantibody negative, no insulin treatment for >3 years post diagnosis). C-peptide measurements are available on a subset of individuals in DARE. *median 96 (IQR 36,149) months diabetes duration,

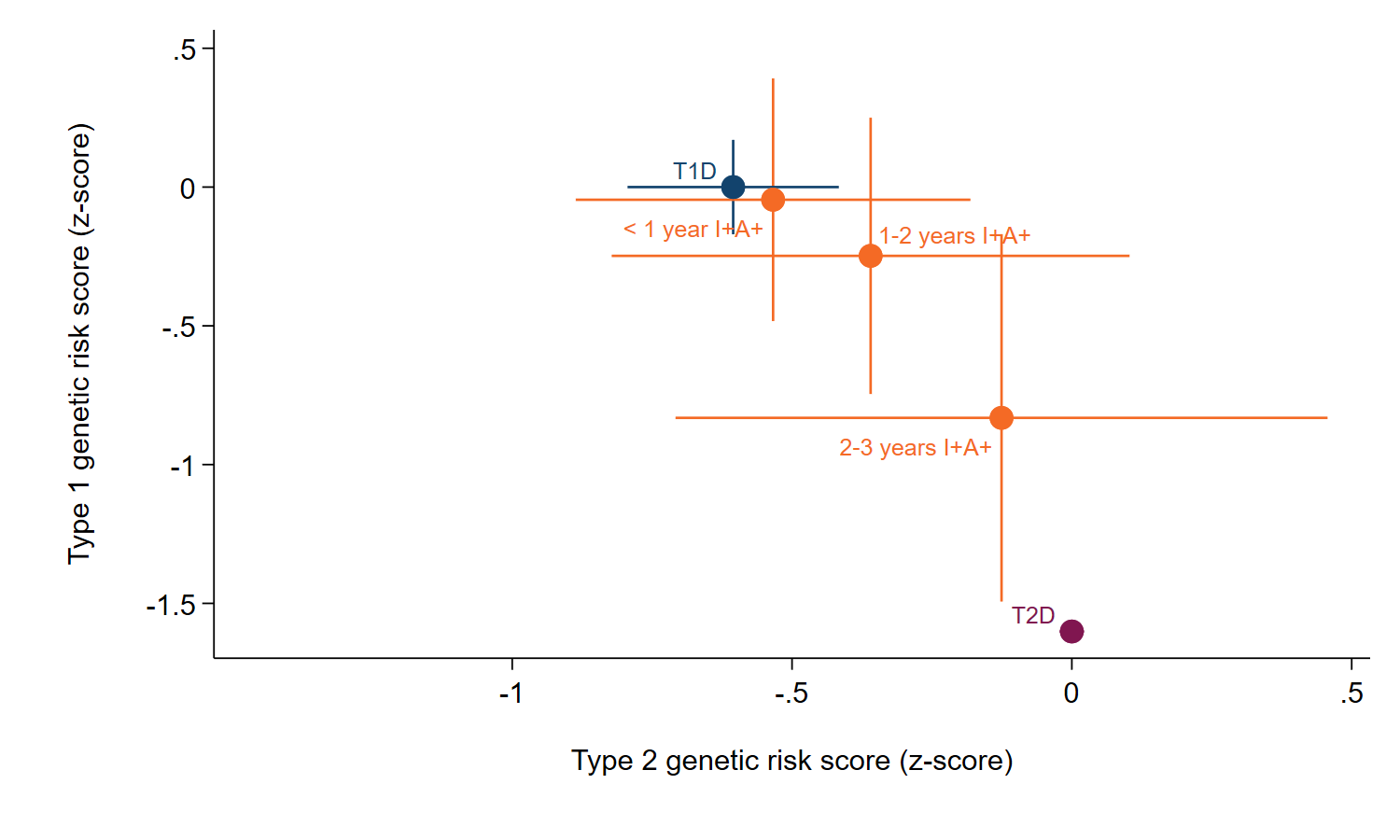
Figure 8s: Genetic risk score for antibody positive individuals in the DARE/GoDARTS cohort who progress to insulin, split by time to insulin, compared with a reference group of individuals with type 1 diabetes and type 2 diabetes. Progression to insulin (I+, time to insulin in years), islet autoantibody status (A+). Error bars represent 95% confidence interval.

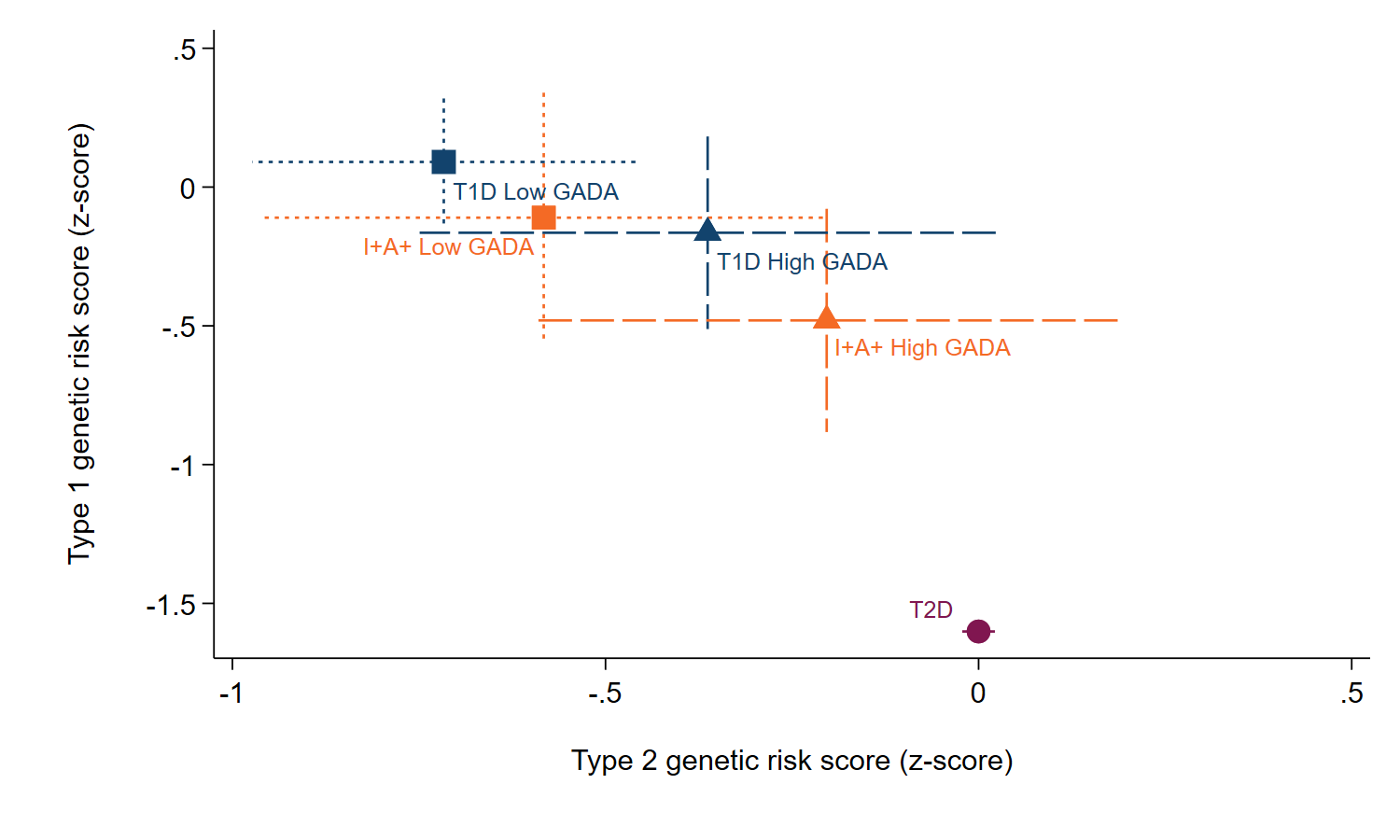

Figure 9s: Genetic risk score for individuals in DARE/GoDARTS split by low (square) and high (triangle) GADA titre compared with a reference group of individuals with type 1 diabetes (dark blue) and type 2 diabetes (purple). Error bars represent 95% confidence interval.
